# *De novo* variants in *LDB1* are linked to distinct neurodevelopmental phenotypes determined by variant location and differing pathomechanisms

**DOI:** 10.64898/2026.02.26.26347174

**Authors:** Rebecca Fluri, Mireia Coll-Tané, Theresa Brunet, Benjamin Cogne, Solene Conrad, Mathilde Nizon, Francesco Nicita, Lorena Travaglini, Antonio Novelli, Margie Glissmeyer, Amanda Peterson, Jillian G. Buchan, Dan Serber, Kolja Meier, Jutta Gärtner, Susann Diegmann, Veronique Pingault, Tania Attie-Bitach, Thomas Courtin, Michael C. Schneider, Wing Hung, Inderneel Sahai, Lauren O’Grady, Katharina Steindl, Sarju G. Mehta, Christel Depienne, Delphine Heron, Boris Keren, Solveig Heide, Shane McKee, Franco Laccone, Lisa M. Dyer, Catherine Melver, Connie Motter, Wendy D. Jones, Zoey Trueblood Wilson, Divya Vats, Kristina Huß, Christiane Zweier, Heinrich Sticht, Anne Gregor

**Affiliations:** Department of Human Genetics, Inselspital University Hospital Bern, University of Bern, 3010 Bern, Switzerland; Department for Biomedical Research (DBMR), University of Bern, 3010 Bern, Switzerland; Department of Human Genetics, Radboud University Medical Center, 6525 GA Nijmegen, the Netherlands; Donders Institute for Brain, Cognition and Behaviour, Radboud University Medical Center, 6525 GA Nijmegen, the Netherlands; Technical University of Munich, TUM School of Medicine and Health, Institute of Human Genetics, TUM University Hospital, 81675 Munich, Germany; Nantes Université, CHU de Nantes, Service de Génétique médicale, F-44000 Nantes, France; Nantes Université, CHU de Nantes, CNRS, INSERM, l’institut du thorax, F-44000 Nantes, France; Unit of Muscular and Neurodegenerative Diseases, Bambino Gesù Children’s Hospital, IRCCS, 00165 Rome, Italy; Laboratory of Medical Genetics, Translational Cytogenomics Research Unit, Bambino Gesù Children’s Hospital, IRCCS, 00165 Rome, Italy; Seattle Children’s Hospital, Seattle, WA 98105, USA; Department of Molecular and Medical Genetics, Oregon Health and Science University, Portland, OR 97239, USA; Department of Laboratory Medicine and Pathology, University of Washington, Seattle, WA 98195, USA; Department of Pediatrics and Adolescent Medicine, Division Pediatric Neurology and German Center for Child and Adolescent Health (DZKJ), University Medical Center Göttingen, 37075 Göttingen, Germany; Service de Médecine Génomique des maladies rares, AP-HP. Centre, Hôpital Necker-Enfants Malades, 75015 Paris, France; Center for Molecular and Chromosomal Genetics, AP-HP-Sorbonne University, Pitié-Salpêtrière Hospital, 75013 Paris, France; Section of Neurology, Department of Pediatrics, St. Christopher’s Hospital for Children, Drexel University College of Medicine, Philadelphia, PA 19134, USA; Division of Medical Genetics and Metabolism, Massachusetts General Hospital for Children, Boston, MA 02114, USA; Institute of Medical Genetics, University of Zurich, 8952 Zurich, Switzerland; Department of Clinical Genetics, Cambridge University Hospitals NHS Foundation Trust, and University of Cambridge, Cambridge CB2 0QQ, UK; Institute of Human Genetics, University Hospital Essen, University of Duisburg-Essen, 45122 Essen, Germany; Département de Génétique, AP-HP-Sorbonne Université, Hôpital Trousseau & Groupe Hospitalier Pitié-Salpêtrière, 75013 Paris, France; Northern Ireland Regional Genetics Service, Belfast City Hospital, Belfast BT9 7AB, UK; Center for Pathobiochemistry and Genetics, Institute of Medical Genetics, Medical University of Vienna, 1090 Vienna, Austria; GeneDx, LLC, Gaithersburg, MD 20877, USA; Division of Medical Genetics, Akron Children’s Hospital, Akron, OH 44302, USA; The North East Thames Regional Genetics Service, Great Ormond Street Hospital, London WC1N 3JH, UK; Southern California Kaiser Permanente Regional Metabolic Genetics Center, Los Angeles CA 90041, USA; LMU Klinikum, Campus Innenstadt, Dr. von Haunersches Kinderspital und iSPZ Hauner MUC, 80337 München, Germany; Institut für Biochemie, Friedrich-Alexander-Universität Erlangen-Nürnberg, 91054 Erlangen, Germany

## Abstract

*LDB1* encodes transcriptional regulator protein LIM-domain-binding protein 1, which plays an important role in neurogenesis. Few C-terminal likely gene disrupting (LGD) variants have been reported in the literature in individuals with congenital ventriculomegaly. Through international collaboration, we now assembled a cohort of 16 individuals with *de novo* variants affecting various regions of *LDB1.* Eleven variants affect either the whole gene or the N-terminal dimerization domain (including gene deletions, NMD-sensitive LGD-, and missense variants) and five variants (missense or NMD-escaping LGD variants) affect only the C-terminus of LDB1 containing the LIM interaction domain. All individuals showed variable neurodevelopmental phenotypes, including developmental delay and behavioral anomalies. In line with literature reports, individuals harboring C-terminal variants additionally presented with ventriculomegaly, establishing a genotype-phenotype correlation. In accordance, we found diverging pathomechanisms *in vitro*: N-terminal missense variants disrupt homodimerization of LDB1, likely leading to a loss of function, while C-terminal variants impair interaction with the essential partner LHX2 in a dominant-negative manner. These findings were confirmed *in vivo* in *Drosophila melanogaster*. Toxicity of overexpressed human LDB1 in *Drosophila* was not observed with N-terminal missense variants but was exacerbated by C-terminal variants. Similarly, phenotypes associated with *LDB1/chi* loss were rescued by overexpression of wild-type LDB1, but neither by LDB1 harboring N-terminal missense variants nor by C-terminal variants that even worsened phenotypes. In summary, our findings link *de novo* variants in *LDB1* to two overlapping, but distinct neurodevelopmental phenotypes based on variant location, and highlight two separate pathomechanisms underlying *LDB1*-related neurodevelopmental disorders.

## Introduction

The *LIM domain binding gene 1 (LDB1*, also known as *NLI* and *CLIM2)* (MIM: 603451) encodes a ubiquitously expressed co-regulator of transcription.^1^ Studies in mice have shown its importance in neurogenesis including development of the fore-,^1^ mid- and hindbrain.^2^ Additionally, it plays an essential role in the differentiation of corticospinal motoneurons and interneurons,^3^ axon guidance,^4^ retinal gliogenesis^5^ and expression of receptors in olfactory sensory neurons.^6^ Its effects outside the nervous system are diverse and notably include regulation of hematopoiesis, cardiogenesis and metabolic functions in hepatocytes and pancreatic cells.^7^

LDB1 itself is a non-DNA-binding protein that regulates transcription through the formation of multimeric protein complexes. Depending on its interaction partners it can act as a transcriptional activator or repressor. It has two domains that are essential for protein-protein interactions: the N-terminal dimerization domain (DD) enables homodimerization and the C-terminal LIM interaction domain (LID) facilitates heterodimerization with LIM proteins, including LIM domain only (LMO) and LIM homeobox proteins (LHX).^8,9^ LIM proteins play an important role in embryonic development and cell type determination. Disruptions of various LHX proteins in different models have revealed their importance specifically in neuronal development.^8^ *De novo* variants in the partner protein *LHX2* (MIM: 603759) have recently been identified as a cause of a neurodevelopmental disorder (NDD),^10^ with one of the described disease mechanisms being an impaired interaction capability with LDB1.^10^ Very recently, also *LDB1* has been tentatively linked to an NDD. Eight C-terminal likely gene disrupting (LGD) variants and one missense variant have been described in individuals with an NDD mainly characterized by congenital ventriculomegaly.^11–13^ Other, less consistently observed clinical features included global developmental delay, dysmorphic features and autism in some cases. However, individual level clinical data are missing for the majority of cases.^11–13^

To better understand the clinical presentation of *LDB1*-associated NDDs, we established a cohort of 16 individuals with variable neurodevelopmental phenotypes, harboring novel missense and LGD variants distributed across *LDB1*. In functional assays, we observed alterations in protein expression levels and impaired protein-protein interactions depending on variant location. Rescue models and overexpression of mutant LDB1 in *Drosophila melanogaster* further corroborated different pathomechanisms based on variant location, indicating a loss-of-function effect for N-terminal variants and a dominant-negative effect for C-terminal variants.

## Subjects and Methods

### Subjects

Through personal communication, GeneMatcher^14^ and DECIPHER,^15^ information on 16 individuals with variants in *LDB1* was assembled. Clinical data were collected using a standardized excel spreadsheet (Table S1). Testing by chromosomal microarray, exome or genome sequencing was done either in a diagnostic setting without the need for specific ethical approval (“clinical”) or in a research setting with ethical approval from the respective review boards (see Table S1). Variants were annotated according to the MANE select transcript (NM_001113407.3; NP_001106878.1) and preliminarily classified according to ACMG criteria^16^ plus current ClinGen sequence variant interpretation recommendations (https://clinicalgenome.org/tools/clingen-variant-classification-guidance/). The criteria application and variant classification presented in this paper does not necessarily reflect the criteria and classification of the clinical testing laboratories involved in this paper.

The individuals, their parents or legal guardians have given informed consent to the publication of the data and pictures. The study complied with the principles set out in the Declaration of Helsinki.

### In-silico analysis

Multiple sequence alignment for conservation analysis of missense variants was done using Clustalw2.^17^ Pathogenicity for missense variants was assessed using different prediction tools: CADD,^18^ REVEL,^19^ SIFT,^20^ AlphaMissense,^21^ Primate AI.^22^ Potential effects on splicing were assessed using SpliceAI.^23^ Disorderedness of C-terminal LID-affecting variants was assessed using PONDR (https://www.pondr.com/). Charge analysis was performed using the EMBOSS charge tool with a window size of 8 (https://www.bioinformatics.nl/cgi-bin/emboss/charge). Isoelectric points (pI) of wild type and mutated proteins were calculated using the Expasy compute pI tool (https://web.expasy.org/compute_pi/).

Variants p.(Arg121Trp), p.(Arg181Gln) and p.(Arg193Trp) were modelled based on the crystal structure of the LDB1 dimerization domain (PDB: 8HIB;^24^). For the interpretation of the p.(Thr359Pro) variant, the LDB1-LHX2 complex was modelled using the structure of the homologous LDB1-LHX3 complex (PDB: 2JTN;^25^) as a template. Variant residues were introduced in the wild-type structures using SwissModel^26^, and RasMol^27^ was used for structure analysis and visualization.

### Plasmids

Expression constructs containing N-terminally FLAG- or HA-tagged *LDB1* (isoform: NM_001113407.3) were subcloned into the pcDNA3.1 vector. Four novel missense variants [p.(Arg121Trp), p.(Arg181Gln), p.(Arg193Trp) and p.(Thr359Pro)] and six novel or previously reported C-terminal LID-disrupting variants [novel: p.(Ser317Argfs*21), p.(Gln333*), p.(Arg356Serfs*127); previously reported: p.(Gly304Glnfs*22), p.(Ser319Argfs*18), p.(Glu349Serfs*134)] were introduced into the FLAG-LDB1 vector via site-directed mutagenesis using a modified version of the Quikchange site-directed mutagenesis kit (Stratagene, Agilent, Santa Clara, CA, USA). A construct containing Myc-tagged *LHX2* (isoform: NM_004789.4) was generated previously.^10^

### Protein expression analysis

For protein expression analysis HEK239 cells were transiently transfected with FLAG-tagged wild-type or mutant *LDB1* (1.5µg plasmid per 6 well) using the jetPrime system (Polyplus transfection, Illkirch France). One to two days after transfection, cells were lysed with RIPA-Buffer (50mM Tris pH 8.0, 150mM NaCl, 1% Igepal CA-630, 0.1% SDS, 0.5% Sodium-deoxycholate). To assess protein stability, cells were treated with 25µM MG132 (Sigma Aldrich, St. Louis, MO, USA) 24 hours post transfection and incubated at 37°C for 4h prior to lysis.

For SDS-Page, protein lysates were mixed with 4x SDS-Page sample buffer, separated on a 4-20% Mini-PROTEAN TGX Stain-Free gel (Bio-Rad, Hercules, CA, USA) and subsequently blotted onto a nitrocellulose membrane using the semi dry blotting system (Bio-Rad). Membranes were incubated with the primary antibodies rabbit anti-FLAG (F7425, Sigma, 1:5,000) and rabbit anti-Histone H3 (4499, Cell Signaling Technology (CST), Danvers, MA, USA, 1:5,000), followed by incubation with goat anti-rabbit HRP conjugated secondary antibody (170-6515, Bio-Rad, 1:15,000). SuperSignal West Femto Maximum Sensitivity Substrate (Thermo Scientific, Waltham, MA, USA) was used to visualize proteins with the ChemiDoc Imaging System (Bio-Rad). Analysis was done with Image Lab software version 6.1 (Bio-Rad). Experiments were carried out at least in triplicates. Protein expression of LDB1 was normalized to the corresponding expression of Histone H3 and normalized expression values were log_2_ transformed. Statistical significance was assessed using a one sample *t*-test with theoretical test value set to 0. An overview of all antibodies used can be found in Table S2.

### Immunofluorescence analysis

HeLa cells seeded on coverslips were transfected with wild-type or mutant LDB1 (500ng per 12 well) using the jetPrime system. After 48h, the cells were fixated with 4% paraformaldehyde in phosphate-buffered saline (PBS) and permeabilized with 0.1% Triton X-100 in PBS. They were stained with primary antibodies mouse anti-FLAG (F1804, Sigma Aldrich, 1:250) and rabbit anti-Nucleolin (14574, CST, 1:100) and the secondary antibodies Alexa Fluor 488 goat anti-rabbit (A11008, Thermo Scientific, 1:500) and Alexa Fluor 546 donkey anti-mouse (A10036, Thermo Scientific, 1:500), nuclei were counterstained with DAPI (Serva, Heidelberg, Germany, 1:50’000). The Zeiss Axio Imager Z2 with Apotome3 (Carl Zeiss, Oberkochen, Germany) with a 63x objective and Zen software v3.13 was used for imaging and analysis.

### Co-Immunoprecipitation

To assess the interaction capability of LDB1 with interaction partners, HEK 293 cells were transfected with FLAG-tagged wild-type or mutant *LDB1* (1.5µg per 6 well) alongside either Myc-tagged *LHX2* or HA-tagged wild-type *LDB1* (0.5µg per 6 well). Additionally, *LDB1* constructs with C-terminal LID-affecting variants were transfected together with wild-type *LDB1* (0.75µg mutant and 0.75µg wild-type *LDB1* per 6 well) and *LHX2*. Two days after transfection the cells were lysed with RIPA-buffer and the extracted proteins incubated with anti-FLAG M2 magnetic beads (Sigma Aldrich) rotating overnight. Beads were washed with RIPA buffer and Tris-buffered saline (TBS), and proteins were subsequently eluted from the beads in 1x SDS-Page sample buffer. SDS-PAGE of samples was performed as described for expression analysis above. Primary antibodies rabbit anti-HA (3724, CST, 1:2000), rabbit anti-Myc (2272S, CST, 1:2500) and rabbit anti-FLAG (1:5000) and secondary antibody goat anti-rabbit HRP (1:15000) were used. Experiments were replicated at least three times. For quantification, IP of Myc-LHX2 or HA-LDB1, respectively, was normalized to the corresponding IP band(s) of FLAG-LDB1 and log_2_ transformed. A one sample *t*-test with theoretical value set to 0 was used to calculate the p-values.

### *Drosophila* stocks and maintenance

*Drosophila melanogaster* stocks were raised at room temperature in clear vials containing fly food (agar, yeast, corn flour, sugar, 0.1% methyl paraben and propionic acid) and sealed with mite-proof stoppers. To induce tissue specific knockdown/overexpression the UAS/GAL4 system was used.^28^ All crosses were carried out at 28°C to increase knockdown/overexpression efficiency.^29^

Stocks were obtained from the Bloomington Drosophila Stock Center [BDSC; pan-neuronal drivers elav-GAL4/CyO (BL#8765) and elav-GAL4 (BL#8760), glial driver repo-GAL4/Tm3 Sb (BL#7415), ubiquitous drivers actin-GAL4/Tm3 Sb Tb (BL#3954) and actin-GAL4/CyO (BL#4414), motoneuron-specific driver D42-GAL4 (BL#8816), RNAi 2 (BL#31049), RNAi 3 (BL#35435), control 2 (BL#36303), OE chi (BL#67741), control hOE (#BL24749)] and the Vienna Drosophila research center [VDRC; RNAi 1 (vdrc107314/KK), control 1 (vdrc60100), RNAi 4 (vdrc30454), control OE (vdrc60000)] or assembled/re-balanced in-house (glial driver repo-GAL4/Tm6 Sb Tb).

To generate UAS-lines for overexpression of human wild-type and mutant LDB1, the coding sequence of *LDB1* was cloned into the pUAST-attb vector (DGRC Stock 1419, https://dgrc.bio.indiana.edu//stock/1419). Patient-specific variants were introduced using site-directed mutagenesis [p.(Arg121Trp), p.(Arg181Gln), p.(Arg193Thr), p.(Thr359Pro), p.(Ser317Argfs*21)]. After sequence verification, constructs were sent to FlyORF (Zurich, Switzerland) for injection into the line BDSC #24749 (control hOE) to create transgenic flies. For rescue experiments, double transgenic flies simultaneously expressing RNAi 1 and either wild-type or mutant LDB1 were generated using a double balancer line (Kr/CyO;D/Tm6c Sb Tb). See Table S3 for an overview of all fly lines.

### RNA isolation and expression analysis

To assess the knockdown/overexpression efficiency of *LDB1* and its *Drosophila* ortholog *chi*, crosses with ubiquitous dosage manipulation of *chi/LDB1* using the actin-GAL4/Tm3 Sb Tb driver and respective *chi/LDB1*/control lines were carried out. Non tubby larvae of the progeny were collected, and RNA was isolated using a modified RNeasy Lipid Tissue Mini Plus Kit (Qiagen, Germantown, MD, USA), where Qiazol was replaced with TRIzol (Thermo Scientific). To perform expression analysis, RNA was converted into cDNA using the Superscript II Reverse Transcriptase and random hexamer primers (Thermo Scientific). Quantitative RT-PCR reaction was run on a QuantStudio3 using PowerTrack SYBR Green Mastermix (Thermo Scientific) and two primer pairs to amplify *chi* (F1:CAACGACCACCCAACAAGAG / R1:TCGTCCTCCTCACCAAACTC and F2:GAACATTCCCGGCAACTACC / R2:CTGGCCACTGTTAAACGGAG) or a primer pair to amplify *LDB1* (F:TGTCACGCCACAAGACCTAC / R:CTGACATCTTCCGTTTCCGC). The subsequent analysis was done using the QuantStudio Design and Analysis software 2.5.1 (Thermo Scientific), and the relative expression of *chi/LDB1* was determined using the ΔΔCT method with tubulin as endogenous control.

### Viability analysis

Viability was assessed for fly lines with ubiquitous dosage manipulation of *chi* and fly lines with ubiquitous overexpression of mutant or wild-type LDB1 in a *chi*-deficient or wild-type background. Two actin-GAL4 driver lines (actin-GAL4/Tm3 Sb Tb and actin-GAL4/CyO) were used for dosage manipulation. Crosses were set up in duplicate vials, and flies were counted separated by sex and genotype daily for 4-6 days until all flies had hatched. Results were confirmed in at least one additional independent experiment. Statistical significance was determined by comparing the percentage of actin-GAL4 vs. Balancer (Tm3 Sb Tb or CyO) flies using a student’s t-test with Bonferroni correction to account for multiple comparisons.

### Negative geotaxis assay

Climbing behavior was assessed using the negative geotaxis assay^30^ and performed as described elsewhere.^31^ Dosage manipulation was induced either pan-neuronally (elav-GAL4/CyO), motoneuron-specific (D42-GAL4) or in glial cells (repo-GAL4/Tm3 Sb). In brief, flies were collected 0-48h post eclosion and transferred into vials, each containing 5 female and 5 male flies. At least 10 vials with 10 flies (total = 100 flies) were tested per condition/genotype, and the results were validated in at least one independent experiment. After recovery from CO_2_ anesthesia for at least 24h, flies were transferred to test vials with an adjustment time of 1 min. The vials were placed below a light source, flies tapped to the bottom of the vial and filmed with a camera (Canon PowerShot, SX620 HS). From the film, the fraction of flies that climbed above the target line at 8 cm after 10s were measured using the software avidemux. The examiner was blinded to the genotype of the flies during the analysis. Statistical analysis was done using the Wilcoxon signed rank test in Rstudio (V2022.07.0x548) with Bonferroni correction to account for multiple comparisons.

### Bang sensitivity assay

Seizure susceptibility was assessed using the bang sensitivity assay as previously described.^31^ Dosage manipulation was induced either pan-neuronally (elav-GAL4/CyO) or in glial cells (repo-GAL4/Tm3 Sb). Fly collection was done as described for the climbing assay. The assay was performed by vortexing the flies for 10s and subsequently measuring the fraction of flies undergoing spasms 5s after being lifted from the vortex (Vortex Genie 2, Scientific Industries). The filming, analysis and statistical analysis were done with the same tools as the negative geotaxis assay.

### Sleep monitoring and analysis

Locomotor activity and sleep were recorded with the Trikinetics *Drosophila* Activity Monitor (DAM2) system (Waltham, MA, USA). Dosage manipulation was induced either pan-neuronally (elav-GAL4), or in glial cells (repo-GAL4/Tm6 Sb Tb). In brief, 3- to 5-day-old male flies were individually placed without CO_2_ anesthesia in transparent tubes (65 mm x 5 mm) containing standard food and loaded into the DAM systems. Flies were allowed to acclimate to activity monitors and food for at least 12 hours, and were then monitored for four days at 25°C in a 12:12 light/dark (LD) cycle. Motion was detected via the monitors’ infrared light beams, and sleep (defined in *Drosophila* as five or more minutes of inactivity)^32,33^ parameters were extracted using the publicly available Sleep and Circadian Analysis MATLAB Program (SCAMP)^34^ for MATLAB. The sleep data presented are the average for the four days of data acquisition and from at least three independent experiments (N=3) unless specified otherwise. To assess significance between two genotypes, we used two-tailed unpaired t-tests for data following a Gaussian distribution or Mann-Whitney tests for non-parametric data. For groups of more than two genotypes, one-way ANOVA with Bonferroni correction for datasets following a normal distribution or Kruskal-Wallis test with Dunn’s multiple comparisons for non-parametric data test was performed. Post hoc Bonferroni correction for multiple testing was further applied for the number of tests performed on a dataset per genotype to determine the corrected two-sided significance level. Only p-values that passed the corrected significance level are indicated in the figures. Statistical analysis was carried out in GraphPad Prism version 10 for Windows (GraphPad Software, San Diego, CA, USA).

## Results

### *LDB1*-associated variant spectrum

We assembled a total of 16 variants in *LDB1*, including two small chromosomal deletions encompassing *LDB1* and five N-terminal LGD variants, three of which are nonsense variants [p.(Gln66*), p.(Trp197*), p.(Gln203*)], two frameshift variants [p.(Ala136Cysfs*9), p.(p.(Thr67Leufs*38))/p.(p.(Gln66Lysfs*39) (both occurred mosaic in one individual, predicted to add up to around 50% of reads)] and one canonical splice-site variant [c.(352+1G>A)]. Additionally, we collected four C-terminal LID-disrupting LGD variants [p.(Ser317Argfs*21), p.(Gln333*), p.(Glu349Serfs*134), p.(Arg356Serfs*127)], and four missense variants [p.(Arg121Trp), p.(Arg181Gln), p.(Arg193Trp), p.(Thr359Pro)] (Figure 1A). One of the variants [p.(Glu349Serfs*134)] has previously been published,^12,13^ and we are providing additional clinical details here. All variants with available parental samples for segregation analysis (15/15) occurred *de novo*.

**Figure 1.**
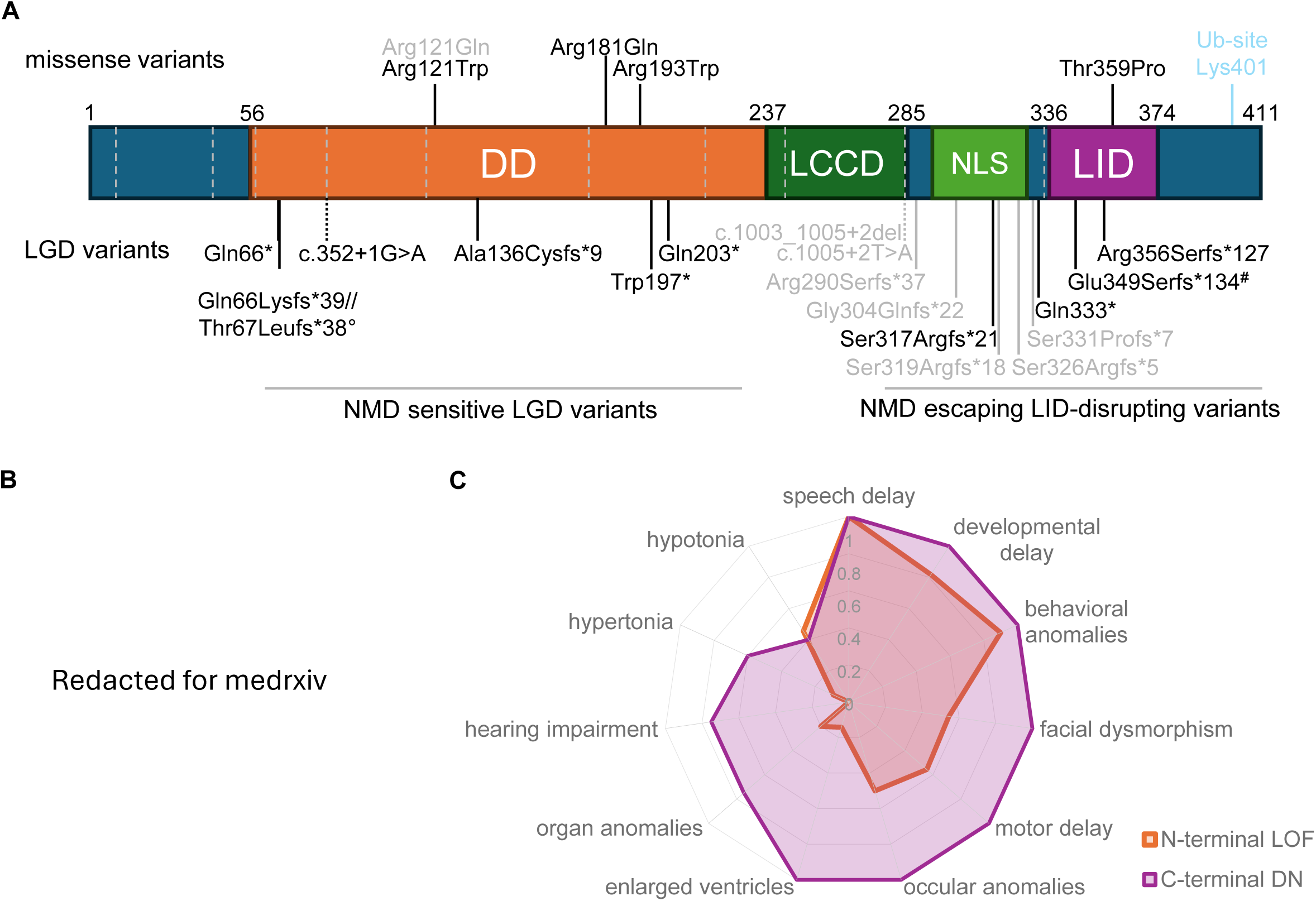
Overview of LDB1 protein structure, identified variants and clinical images of affected individuals. (A) Schematic drawing of LDB1 (NP_001106878.1) with variants assembled in this study labelled in black, and previously published variants^11–13^ in grey. Of note, nomenclature of variants from Allington et al.^13^ was adjusted to correspond with HGSV guidelines. Domains are color-coded and identified according to Wang et al.^9^ Corresponding exon boundaries are highlighted with grey dashed lines. DD – dimerization domain, LCCD – LDB/Chip conserved domain, NLS – nuclear localization signal, LID – LIM interaction domain. Ub – Ubiquitination. °Both variants occur in mosaic form in one individual and are predicted to add up to about 50% of reads, correlating with a heterozygous state. ^#^Variant was previously published,^12,13^ but is here included with additional clinical information on the affected individual. (B) Clinical images of several affected individuals show facial dysmorphism and highlight similarities between cases. (C) Phenotypic differences and similarities between newly and previously reported individuals^11–13^ harboring N-terminal LGD or DD-missense variants (N-terminal LOF) and C-terminal LID-affecting (LID-missense, C-terminal frameshift/nonsense/splice site) variants with dominant negative effects (DN) (C-terminal DN) are shown in a radar plot.

Both chromosomal deletions encompass *PPRC1* (MIM: 617462) alongside *LDB1* with the larger deletion additionally comprising *HPS6* (MIM: 607522) and *ARMH3* (MIM: 620867) and the smaller deletion comprising *NOLC1* (MIM: 602394). None of these genes are associated with a dominant rare disorder to date. N-terminal LGD variants are likely to undergo nonsense mediated mRNA decay (NMD) and subsequently lead to loss of function. In contrast, all C-terminal LGD variants result in a stop codon in the penultimate or last exon and therefore likely escape NMD. These variants are either located shortly before or within the LIM binding domain (LID) of *LDB1*. Most of these C-terminal LID-disrupting variants remove the relatively acidic and disordered C-terminus of LDB1 and consequently increase rigidity of the region and the isoelectric point of the total protein (Figure S1A-C).

Regarding missense variants, one of them [p.(Thr359Pro)] is also situated in the LIM interaction domain (LID), while the other three reside in the dimerization domain (DD). All identified missense variants affect highly conserved amino acids (Figure S1D). Different prediction tools estimate a high chance of pathogenicity for all missense variants (Table S4). The variant p.(Arg181Gln), however, is predicted to be less likely deleterious than the others, having the lowest REVEL (0.53) and AlphaMissense (0.59) scores. The LGD variant c.(352+1G>A) affects a consensus splice site with a high probability of resulting in donor loss (SpliceAI score 0.82), likely leading to skipping of exon 5 and subsequently frameshifting. All variants are very rare and either absent from gnomAD v4.1.0,^35^ or present only in one [p.(Gln203*): 1/1610904] or very few [p.(Arg181Gln): 4/1613342] individuals.

*LDB1* shows strong constraint against loss-of-function (pLI = 1.00, observed/expected = 0.12(0.07-0.24), LOEUF = 0.24) and missense variation (Missense Z score = 3.49, observed/expected = 0.6(0.55-0.66)) according to population based metrics from gnomAD v4.1.0. Furthermore, dosage-sensitivity prediction scores suggest that LDB1 is intolerant to dosage loss (haploinsufficiency; pHaplo = 0.94) and dosage gains (triplosensitivity; pTriplo = 0.90).

When preliminarily classifying *LDB1* variants according to ACMG criteria, all N-terminal LGD variants and C-terminal LID-domain disrupting variants can, together with our functional data (see below), be classified as likely pathogenic or pathogenic. For the missense variants, three of them can also be classified as likely pathogenic, while the fourth variant p.(Arg181Gln) remains a variant of unknown significance (VUS). Classification details and criteria used can be found in Table S4.

### Clinical Spectrum associated with *LDB1* variants

Most affected individuals for whom detailed clinical information was available show developmental delay (14/16) including speech (13/13) and motor delay (8/13), as well as intellectual disability (8/10). The severity in disability ranges from mild to severe with an inability to speech and walk unaided. Behavioral anomalies (13/14), such as hyperactivity, attentional difficulties and autism are also common. MRI anomalies include ventriculomegaly/enlarged ventricles, cerebellar vermis hypoplasia, hypoplastic hippocampi, and partial agenesis of the corpus callosum and facial nerve. Few (2/13) individuals have been reported having epilepsy. Organ malformations outside the nervous system occurred rather infrequently and included cardiac defects (3/13), renal anomalies (2/13), and intestinal anomalies (1/8). Variable urogenital anomalies occurred in more than half of the cases (7/13). Choanal atresia was observed in two patients. Hearing loss (3/14) and ocular abnormalities (9/14) such as cortical visual impairment, coloboma of the optic nerve, ptosis, hyperopia and strabismus have also been observed. Minor variable skeletal anomalies (11/16) are frequently reported. Non-specific facial dysmorphisms are often noted (11/16), and some individuals share characteristic features (Figure 1B). Growth parameters were not consistently altered and normal for many individuals (abnormal parameters for height, weight or head circumference in 7/15). An overview of clinical findings can be found in Table 1, and detailed clinical information for all individuals can be found in Table S1.

**Table 1.** Summary of available clinical finding in individuals with LDB1 variants (novel and published. ^11–13^ **[novel]).** Diverging phenotypes between genotype groups are highlighted in yellow.

| Clinical finding | N-terminal LGD + DD-missense (n = 12 [11]) | C-terminal LID-affecting (n = 12 [5]) |
| --- | --- | --- |
| Developmental delay | 9/11 [9/11] | 7/7 [5/5] |
| Speech delay | 10/10 [10/10] | 5/5 [3/3] |
| Behavioral anomalies | 9/10 [9/10] | 4/4 [4/4] |
| Facial dysmorphism | 6/11 [6/11] | 6/7 [5/5] |
| Epilepsy | 2/11 [2/11] | 0/2 [0/2] |
| Ventriculomegaly/ enlarged ventricles | 1/7 [0/6] | 9/9 [2/2] |
| Other organ malformations (cardiac, renal, GI) | 2/10 [2/10] | 3/4 [2/3] |
| Hypotonia | 5/11 [5/11] | 2/5 [1/3] |
| Hypertonia | 1/11 [1/11] | 3/5 [2/3] |
| Motor delay | 5/9 [5/9] (mostly mild) | 5/5 [3/3] |
| Vision impairment | 5/10 [5/10] | 4/4 [4/4] |
| Hearing loss | 0/10 [0/10] | 3/4 [3/4] |

### Genotype-phenotype correlations in individuals with *LDB1* variants

When combining data from this cohort with the sparse available data from published cases, tentative genotype-phenotype correlations can be established. As the number of cases is still rather limited, no statistical validation can be performed. Ventriculomegaly is found in all individuals with detailed clinical information and C-terminal LID-disrupting (C-terminal frameshift, nonsense and splice site) variants, but only in one previously published individual with a missense variant in the DD region. Additionally, severe motor delay, hearing loss, eye abnormalities, facial dysmorphism and involvement of non-neuronal organs (e.g. heart, kidney, GI-tract) tend to be more prevalent in individuals with C-terminal LID-affecting variants (LID-missense and C-terminal LID disrupting variants). Interestingly, if affected muscle tonus is altered, it seems to be mirrored between different mutation types: muscular hypotonia appears to be slightly more prevalent in individuals with N-terminal LGD or missense variants while hypertonia is predominantly present in individuals with C-terminal LID-affecting variants (Figure 1C). A summary of phenotypic differences and similarities between variant groups can be found in Table 1.

### Mutational modelling

Structural analysis of the dimerization domain (DD) revealed that the arginine residues 121, 181, and 193 all form stabilizing polar interactions within the DD (Figure 2A,C,E). In the p.(Arg121Trp) and p.(Arg193Trp) variants, these polar interactions are completely disrupted by the aromatic tryptophan sidechain (Figure 2B,F). Therefore, these variants are predicted to cause severe destabilization of the DD, which is also likely to affect its dimerization properties. In the p.(Arg181Gln) variant, a salt-bridge between the charged wild-type residues Arg181 and Asp158 is replaced by a weaker Gln181-Asp158 hydrogen bond (Figure 2C,D). Since this variant at least preserves a polar interaction, it is predicted to have a weaker effect on protein structure compared to the variants at position 121 and 193.

**Figure 2.**
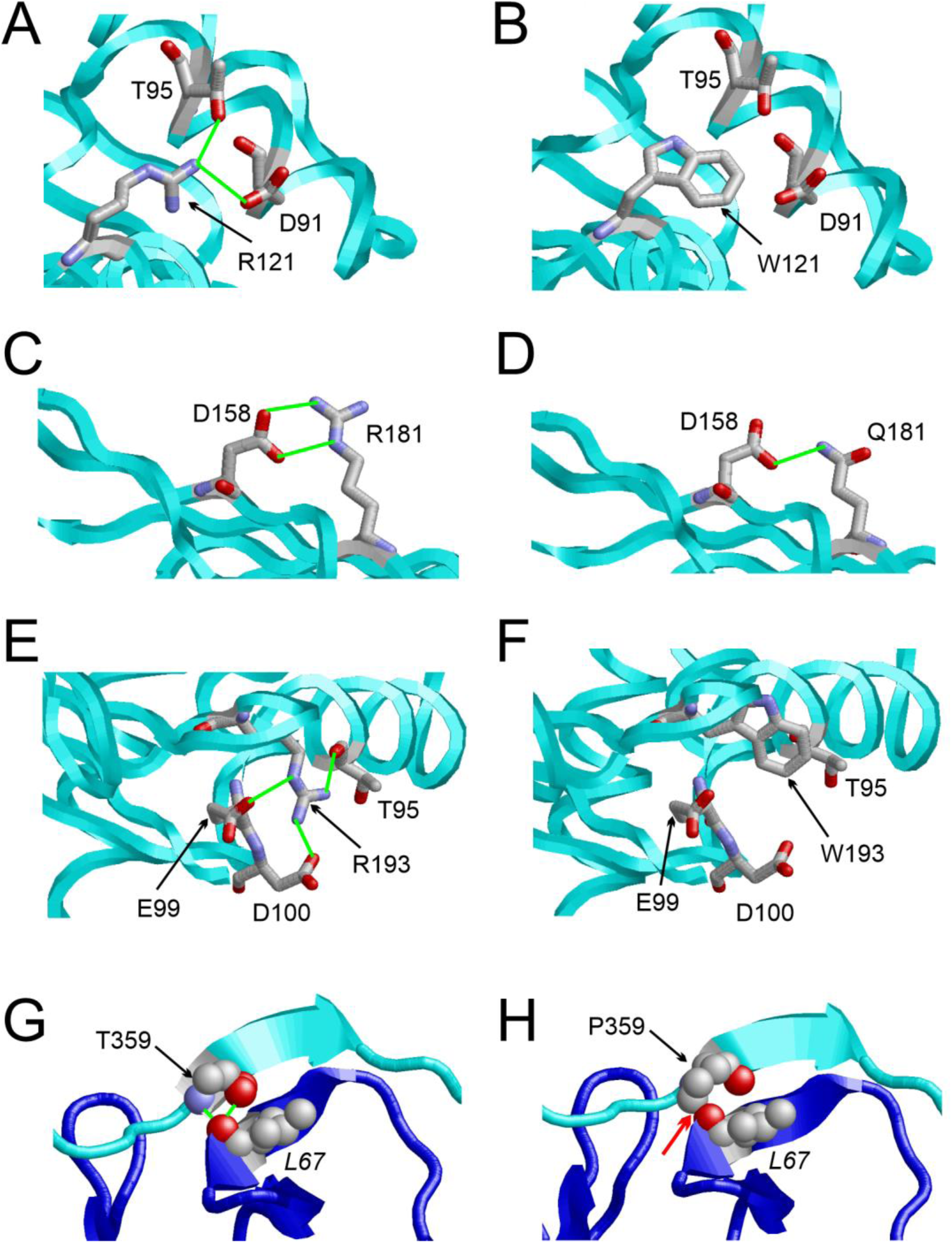
Structural effects of the LDB1 missense variants. (A) Arg121 of the dimerization domain forms polar interactions (green lines) with Asp91 and Thr95. All interacting residues are shown in stick presentation and are colored according to the atom types. The backbone topology of LDB1 is shown as cyan ribbon. (B) The aromatic sidechain of variant Trp121 cannot form stabilizing interactions with Asp91 and Thr95. (C) The charged Arg181 forms a salt-bridge (green lines) with Asp158. (D) The polar but uncharged Gln181 exhibits a reduced interaction with Asp158. (E) Arg193 forms polar interactions (green lines) with Thr95, Glu99, and Asp100. (F) The aromatic sidechain of variant Trp193 cannot form stabilizing interactions with Thr95, Glu99, and Asp100. (G) Thr359 of the LID forms polar interactions (green lines) with Leu67 of LHX2. The interacting residues are shown in ball-and-stick presentation and colored according to the atom types. The backbone of LDB1 and LHX2 is shown as cyan and blue ribbon, respectively. (H) The variant Pro359 cannot form these polar interactions and causes a steric clash (red arrow) with Leu67 instead.

The p.(Thr359Pro) variant is directly located at the interface between the LDB1 LID and LHX2 (Figure 2G,H). Thr359 forms stabilizing polar interactions with Leu67 of LHX2. These interactions cannot be formed by the cyclic and nonpolar Pro359, which additionally causes steric clashes with Leu67. Therefore, this variant is predicted to destabilize the LDB1-LHX2 interface.

### Altered LDB1 expression of missense and C-terminal LID-disrupting variants

To gain a better understanding of the functional effects of variants in *LDB1*, we conducted *in vitro* experiments. All novel and several previously published missense and C-terminal LID-disrupting variants (Figure 3A) were included in the cellular experiments.

**Figure 3.**
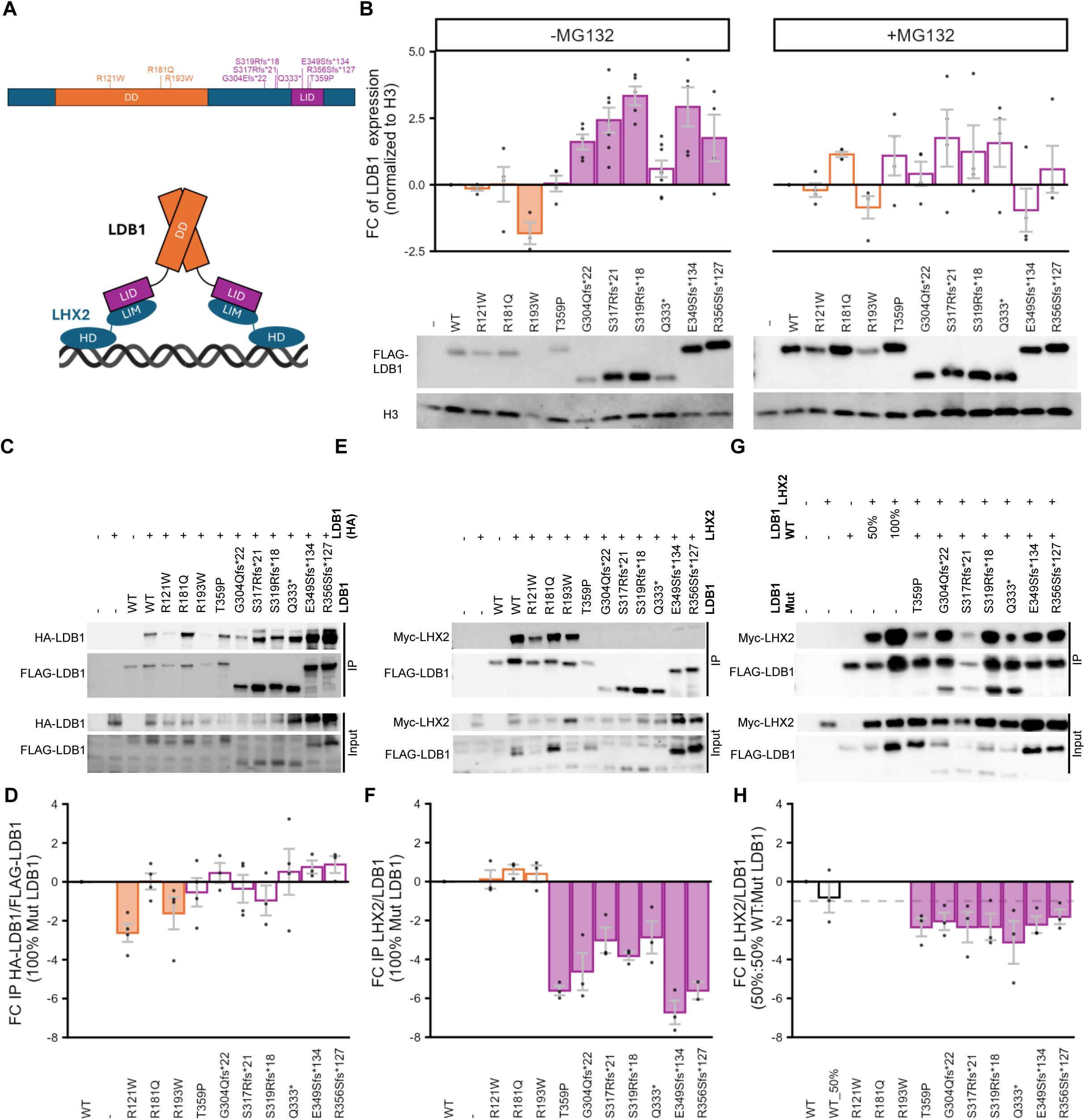
*LDB1* variants affect expression levels and interaction with essential interaction partners. (A) Schematic overview of *LDB1* variants tested in cellular assays and schematic of LDB1-LHX2 tetramer formation together with DNA is shown. Variants are color-coded according to their domain-affectation (orange = dimerization domain (DD), purple = LIM interaction domain (LID)). (B) Expression analysis of *LDB1* transiently transfected in HEK293 cells with (+MG132) and without (-MG132) treatment with proteasomal inhibitor MG132. Without MG132 treatment, reduced expression levels were found for p.(Arg193Trp) and increased expression levels for several C-terminal frameshift variants. A representative image of the Western Blot and quantification from three independent experiments are shown. LDB1 expression levels were normalized to H3. (C-H) Co-immunoprecipitation (Co-IP) of FLAG-tagged wild-type or mutant LDB1 with HA-tagged wild-type LDB1 (C, D) or Myc-tagged LHX2 (E-H) showed impaired homodimerization abilities for two DD missense variants and impaired interaction with LHX2 for all C-terminal LID-affecting variants. For interaction with LHX2, simultaneous overexpression of wild-type and mutant LDB1 indicated dominant-negative effects of tested variants (G-H). Representative images (C, E, G) and quantifications from at least three independent experiments (D, F, H) are shown. Grey dashed line in (H) represents expected value for 50% wild-type input. For quantification, IP of HA-LDB1/Myc-LHX2 was normalized to IP of FLAG-LDB1. For all experiments here (B-H), statistical significance was determined using a 1-sample student’s t-test with a theoretical value of 0. Significant expression changes are highlighted with filled bars (p < 0.05). For visualization, normalized expression values were log_2_-transformed. Individual values are shown as black dots. WT – wild-type.

Using transient overexpression constructs, we investigated the protein expression of wild-type and mutant LDB1 in HEK293 cells. We observed that the C-terminal LID-disrupting frameshift variants [p.(Gly304Glnfs*22), p.(Ser317Argfs*21), p.(Ser319Argfs*18), p.(Glu349Serfs*134), p.(Arg356Serfs*127)] significantly increased the protein expression of altered LDB1 compared to wild-type LDB1 (Figure 3B). Interestingly, expression levels were not increased for the C-terminal nonsense variant p.(Gln333*). Additionally, the N-terminal DD missense variant p.(Arg193Trp) significantly decreased the protein expression of LDB1. The other missense variants [p.(Arg121Trp), p.(Arg181Gln), p.(Thr359Pro)] did not consistently alter LDB1 expression (Figure 3B).

To determine whether the differences in protein expression levels were caused by altered degradation rates, transfected HEK293 cells were treated with proteasomal inhibitor MG132 prior to cell lysis. Expression levels of wild-type and mutant LDB1 were no longer significantly different following proteasomal inhibition (Figure 3B), suggesting that the varying protein expression levels were indeed due to an increased or reduced degradation.

We additionally analyzed subcellular localization of wild-type and mutant LDB1 using immunofluorescence analysis. Wild-type LDB1 and LDB1 carrying any of the tested missense variants were diffusely distributed in the nucleus (Figure S2). C-terminal frameshift variants, but not the C-terminal nonsense variant p.(Gln333*), resulted in the formation of punctate non-nucleolar aggregates in the nucleus (Figure S3).

### Impaired interaction capability of mutant LDB1 with partner proteins

Furthermore, we tested whether variants in LDB1 affect the interaction capability of LDB1 with itself and essential partner proteins. To assess the effect of the variants on the homodimerization potential of LDB1, co-immunoprecipitation of FLAG-tagged LDB1 and HA-tagged wild-type LDB1 was performed. Two of the three missense variants [p.(Arg121Trp) and p.(Arg193Trp)] located in the DD of LDB1, impaired homodimerization, suggesting a loss-of-function mechanism. All variants solely affecting the LID did not impair homodimerization (Figure 3C-D).

To test the interaction capability of mutant LDB1 with LIM proteins, co-immunoprecipitation of LDB1 with the LIM Homeobox 2 (LHX2) protein was performed. We observed that the missense variant p.(Thr359Pro) located in the LID and C-terminal LID-disrupting variants almost completely abolish the interaction of LDB1 with LHX2. The other variants located in the DD did not influence the interaction with LHX2 (Figure 3E-F).

As variants are present in a heterozygous state in affected individuals, we wanted to assess whether this interaction was impaired in a dominant-negative manner. To test our hypothesis, we co-transfected wild-type LDB1 with mutant LDB1 and LHX2. We observed that co-transfection of mutant LDB1 containing one of the C-terminal LID-disrupting variants or missense variant p.(Thr359Pro) negatively affects the heterodimerization of wild-type LDB1 with LHX2, confirming our hypothesis that these variants act in a dominant-negative manner (Figure 3G-H).

### Confirming dosage sensitivity of *LDB1* ortholog *chi* in *Drosophila melanogaster*

To gain a better understanding of the importance of *LDB1* in development, we investigated its function *in vivo* using the model system *Drosophila melanogaster*. In flies, *chi* (chip) is the well-conserved ortholog of human *LDB1* and *LDB2*. Knockdown of the *Drosophila* ortholog presents a valuable model for the heterozygous LGD variants observed in our cohort. Due to the suggested dosage sensitivity of *LDB1*, we tested effects of both knockdown and overexpression of commercially available RNAi and overexpression lines. Validation of four different RNAi knockdown lines confirmed a knockdown efficiency to 51-58% residual levels in two of them (RNAi1 and 2) and 6-fold overexpression (OE chi) upon ubiquitous dosage manipulation (Figure S4A-B). Ubiquitous knockdown of *chi* was lethal, and overexpression significantly reduced viability, highlighting its importance for development and the dosage sensitivity of *LDB1/chi* (Figure S4D-E). RNAi lines 3 and 4 were excluded from further experiments due to insufficient knockdown efficiency and consistent lack of lethality phenotypes (Figure S4D).

To gain more insight into the neurodevelopmental functions of *chi*, we assessed basic locomotor function using the negative geotaxis assay upon dosage manipulation of *chi* in neurons, motoneurons, or glial cells. Knockdown and overexpression of *chi* in all neurons or specifically in motoneurons resulted in significant locomotion defects (Figure 4A and S5A). Climbing impairment was more severe upon pan-neuronal knockdown compared to motoneurons only, and most severe with RNAi 1 (Figure S4A). In contrast, glial knockdown did not affect locomotor behavior, while glial overexpression mildly impaired climbing ability. This suggests a more prominent role of *LDB1/chi* in neurons compared to glial cells.

**Figure 4.**
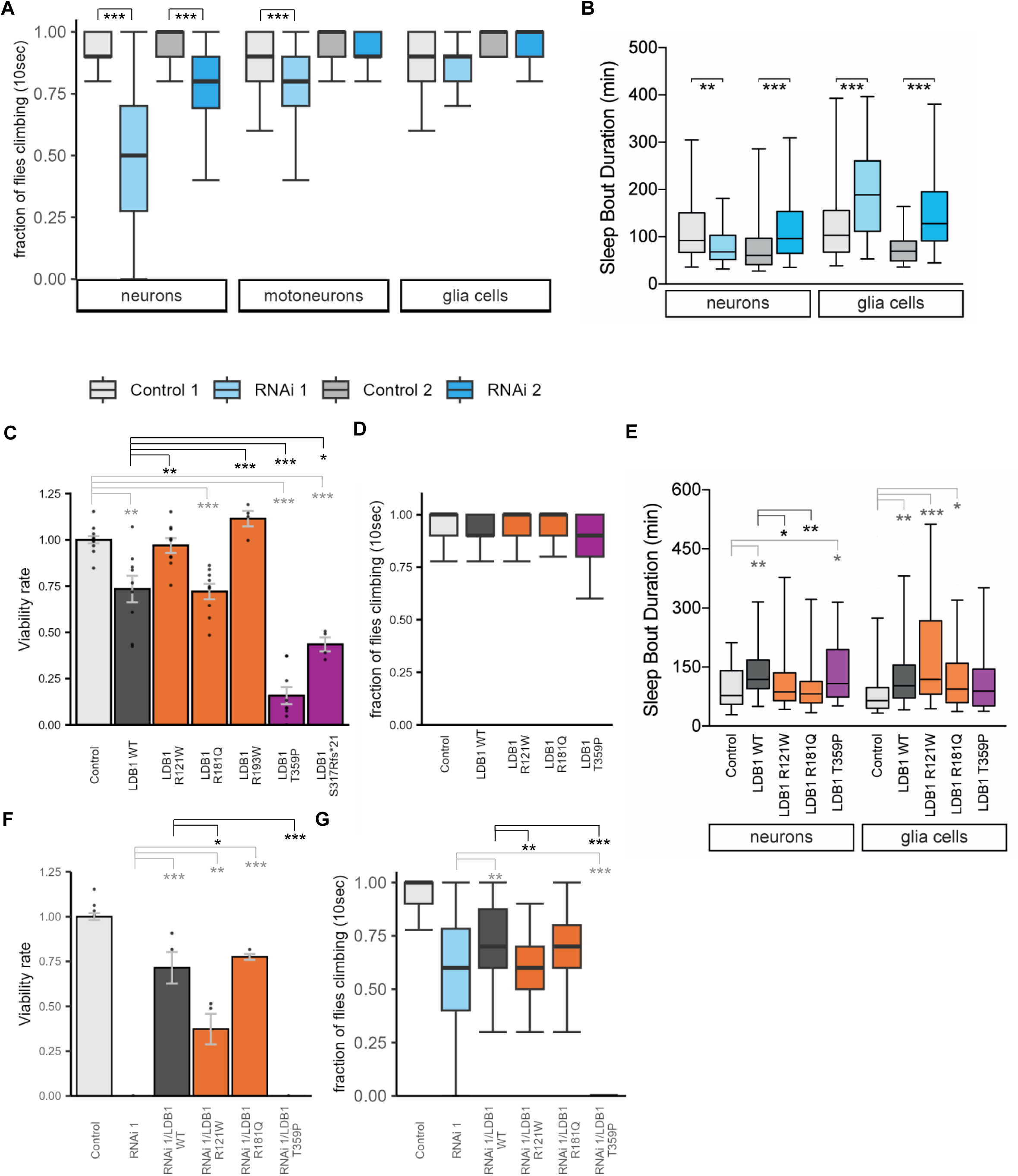
*LDB1/chi* dosage affects viability, negative geotaxis and sleep behavior in Drosophila melanogaster. (A) Negative geotaxis assay showed impaired climbing ability upon pan-neuronal knockdown for two lines and upon motoneuron-specific knockdown of *chi* for one line. Climbing ability was unaffected upon glial knockdown. Fraction of flies climbing at least 8 cm in 10 sec per vial (10 flies per vial) measured. At least 200 flies in batches of ten (n = 20) were tested per genotype. Significance was calculated using a Wilcoxon rank-sum test (*** p < 0.001). (B) Average sleep bout duration during the dark period (ZT12–24). Pan-neuronal *chi* knockdown males showed altered sleep bout duration compared to isogenic controls. Pan-glial *chi* knockdown males exhibited longer sleep bouts than their respective controls. At least 60 flies were tested per genotype. Mann-Whitney or ANOVA tests, with Bonferroni correction for multiple comparisons were performed for statistics. (C) Viability of male flies upon ubiquitous overexpression (using actin-GAL4/Tm3 Sb Tb) of human wild-type of mutant LDB1 was reduced for wild-type and p.(Arg181Gln), and even more strongly reduced for variants p.(Thr359Pro) and p.(Ser317Argfs*21). Viability was normalized to the Control line, which was set to 1. The experiment was carried out independently at least three times in duplicates each. At least 100 flies were counted per experiment and genotype. Significance was calculated on the ratios of non-Sb (overexpression) vs. Sb (Balancer) flies using a student’s t-test with Bonferroni correction for multiple testing. (D) Climbing ability assessed with the negative geotaxis assay was not significantly altered upon pan-neuronal overexpression of wild-type or mutant LDB1. At least 20 vials were tested per genotype. Significance was determined using a Wilcoxon rank-sum test. (E) Average sleep bout duration during the dark period (ZT12–24). Neuronal overexpression of wild-type LDB1 resulted in longer sleep bouts compared to controls. Pan-neuronal overexpression of LDB1 variants p.(Arg121Trp) and p.(Arg181Gln) significantly shortened sleep bouts compared to wild-type overexpression. Glial overexpression of wild-type LDB1 and variants p.(Arg121Trp) and p.(Arg181Gln) led to longer sleep bouts compared to controls. At least 40-70 flies were tested per genotype. For statistical analysis Mann-Whitney or ANOVA tests, with Bonferroni correction for multiple comparisons were done. (F) Ubiquitous Overexpression of human wild-type LDB1 in *chi*-knockdown flies could rescue the lethality phenotype, as could p.(Arg181Gln). Variant p.(Arg121Trp) could rescue the lethality significantly less efficiently, while p.(Thr359Pro) could not rescue at all. The experiment was carried out twice independently in duplicates each with at least 100 flies counted per genotype and experiment. Significance was calculated on the ratios of non-Sb (overexpression) vs. Sb (Balancer) flies using a student’s t-test. (G) Pan-neuronal overexpression of human wild-type LDB1 in neuronal *chi*-knockdown flies could partially rescue climbing impairment observed in the negative geotaxis assay. Variant p.(Arg121Trp) was unable to rescue the defect, while p.(Thr359Pro) significantly worsened the phenotype compared to *chi* knockdown alone. The experiment was carried out twice from independent crossed, and at least n = 20 (200 flies in batches of 10) were tested per experiment. Significance was calculated using a Wilcoxon rank-sum test. Significance: * p < 0.05, ** p < 0.01, *** p < 0.001 (A-G). WT – wild-type. Bars/boxplots are color-coded according to domain-affectation (orange = DD, purple = LID) (C-G). Data are represented as boxplots (25^th^-75^th^ percentiles, median line; whiskers indicate 5^th^-95^th^ percentiles) (A, B, D, E, G) or bar graphs with individual values shown as black dots (C, F).

Knockdown flies did not show any increased seizure susceptibility in the bang sensitivity assay upon neuronal or glial knockdown or overexpression (Figure S5B-C).

Last, we investigated whether *chi* regulates behavior commonly disrupted in neurodevelopmental disorders. Sleep disturbances are highly prevalent in these conditions^36^ and represent a conserved, translatable endpoint in *Drosophila*.^37^ Pan-neuronal knockdown of *chi* using either RNAi line significantly altered sleep episode duration (Figure 4B). However, the direction of the effect differed between knockdown with different RNAi lines: RNAi 1 reduced episode length, whereas RNAi 2 increased it. In the latter case, longer episodes occurred less frequently, indicating enhanced sleep consolidation (Figure S6A). Similarly, glial *chi* knockdown with both RNAi lines consistently led to significantly longer sleep episodes (Figure 4B), accompanied by a reduction in the number of sleep bouts (Figure S6A). These changes suggest that pan-glial *chi* knockdown promotes sleep consolidation and a hypersomnia-like phenotype. Taken together, our findings demonstrate that the *Drosophila* ortholog of LDB1 regulates sleep architecture.

### Overexpression of human LDB1 in *Drosophila melanogaster* leads to toxicity

Having observed different functional consequences of variants in *LDB1* on a cellular level, we tested whether we could observe correlating effects *in vivo* by overexpressing LDB1 in *Drosophila melanogaster*. We first confirmed similar expression levels for wild-type and mutant LDB1 in third instar larvae upon ubiquitous overexpression (Figure S4C). Ubiquitous overexpression (driver: actin-GAL4/Tm3 Sb Tb) of wild-type human LDB1 resulted in reduced viability, indicating toxicity of the human protein (Figure 4C). In contrast, overexpression of mutant LDB1 containing variants p.(Arg121Trp) or p.(Arg193Trp) did not impair viability, indicating a loss-of-function effect. Overexpression of the third DD variant p.(Arg181Gln) impaired viability similar to wild-type LDB1. Additionally, overexpression of LDB1 harboring C-terminal LID-affecting variants p.(Thr359Pro) or p.(Ser317Argfs*21) showed increased toxicity compared to wild-type with an even stronger reduction of viability (Figure 4C). These results were confirmed with a second ubiquitous driver line (actin-GAL4/CyO).

We tested a subset of these variants [p.(Arg121Trp), p.(Arg181Gln), p.(Thr359Pro)] in further assays and rescue experiments. Pan-neuronal overexpression of wild-type or mutant LDB1 did not significantly impair climbing ability in the negative geotaxis assay (Figure 4D). Finally, we assessed the impact of overexpressing LDB1 with any of these variants in neurons or glial cells on sleep architecture. Overexpression of wild-type human LDB1 in neurons resulted in longer sleep bouts (Figure 4E). In contrast, overexpression of LDB1 with the variants p.(Arg121Trp) or p.(Arg181Gln) significantly shortened sleep episodes compared to wild-type LDB1 (Figure 4E). Notably, only flies overexpressing LDB1 with variant p.(Arg181Gln) compensated for this sleep loss by increasing the number of sleep episodes (Figure S6B). In contrast, pan-glial overexpression of wild-type LDB1 or mutant LDB1 with the variants p.(Arg121Trp) or p.(Arg181Gln) significantly increased sleep bout duration (Figure 4E). Interestingly, for p.(Arg121Trp), we observed a tendency toward more consolidated sleep compared to wild-type overexpression.

### Rescue of knockdown phenotype in *chi*-deficient *Drosophila melanogaster* with human LDB1

Expanding on our previous findings, we wanted to test whether overexpression of wild-type or mutant human LDB1 could rescue the *chi* knockdown phenotype observed in *Drosophila melanogaster*. When first looking at ubiquitous knockdown of *chi* (with RNAi 1), additional overexpression of wild-type LDB1 rescued the lethality phenotype and significantly increased viability. Overexpression of mutant LDB1 with the variant p.(Arg181Gln) was similarly effective. In contrast, overexpression of LDB1 with variant p.(Arg121Trp) increased viability by a significantly lesser degree. Furthermore, overexpression of LDB1 with the variant p.(Thr359Pro) did not rescue the knockdown phenotype, also resulting in complete lethality (Figure 4F).

To further substantiate our results, we performed rescue experiments also for the negative geotaxis assay in pan-neuronally *chi*-deficient flies. We observed that overexpression of wild-type LDB1 was partially able to improve the impaired climbing ability caused by *chi* deficiency.

A similar, but not significant rescue tendency was seen for overexpression of LDB1 with the variant p.(Arg181Gln). In contrast, overexpression of mutant LDB1 with the variant p.(Arg121Trp) could not rescue the knockdown phenotype, and overexpression with the variant p.(Thr359Pro) even worsened the basic locomotion defects (Figure 4G).

Together, these findings upon overexpression of wild-type and mutant human LDB1 in control and *chi*-deficient backgrounds support the results from the cellular assays, indicating loss-of-function effects for two of three missense variants from the DD, while variants affecting the LID often lead to more severe phenotypes compatible with dominant-negative effects.

## Discussion

While *de novo* variants in *LDB1* have recently been associated with congenital ventriculomegaly, only a small cohort of mainly C-terminal LGD variants have been studied thus far.^12,13^ By assembling a cohort of 16 individuals with variants in *LDB1* and variable NDD presentations, 15 of which were previously unpublished, we now further expand the clinical phenotype, propose a new genotype-phenotype correlation and highlight diverging pathomechanisms based on variant type and location.

*LDB1*-associated NDD is highly variable with prevalent features including global developmental delay ranging from mild to severe, intellectual disability, behavioral anomalies and non-specific dysmorphic facial features. Congenital ventriculomegaly, the key feature of *LDB1*-associated NDD in the literature,^12,13^ is very frequent in carriers of C-terminal LID-affecting variants in published and novel cases, but absent in the newly described individuals with N-terminal missense- or LGD-variants apart from one previously published case of an N-terminal missense variant.^13^ Additionally, motor delay, hearing loss, ocular anomalies and other non-brain-related organ anomalies were also more frequent in individuals with C-terminal LID-disrupting variants. Though numbers of individuals in each group are still limited, these observations indicate a genotype-phenotype correlation with overlapping but distinct clinical presentations depending on variant type and location. Previous studies have established the importance of LDB1 in embryonic development and neurogenesis. For instance, there is evidence that EMX2 and OTX2 are potential non-LIM partner proteins of LDB1 and involved in the development of the choroid plexus alongside LHX2 and LHX5, providing speculative insight into how LDB1 could be linked to ventriculomegaly.^1,38^

In line with the observed genotype-phenotype correlation, we discovered different functional consequences for N-terminal and C-terminal variants *in vitro* and *in vivo*. All tested C-terminal LID-affecting variants (missense and frameshift/nonsense) impaired heterodimerization of LDB1 with LHX2, and equimolar co-expression of wild-type LDB1 showed that the protein-protein interaction is impaired in a dominant-negative fashion. The LDB1 and LHX2 heterotetramer is critical for various neuronal functions, such as axon guidance and development of the hippocampus.^39^ LHX2-associated NDD shares many similarities with LDB1-associated NDD. Both are highly variable and share key features including developmental delay, intellectual disability, behavioral anomalies and speech impairment. Motor delay, however, was not commonly observed in LHX2-associated NDD, and extra-neural manifestations are even rarer. The LDB1 LIM-interacting domain is highly conserved across species, and its loss impairs interaction with all LIM proteins.^7^ It is therefore likely that the C-terminal variants affect the binding capability of LDB1 with other LHX and LMO as well, potentially resulting in additional features. In *in vivo* experiments, overexpression of C-terminal variants showed even more toxic effects than wild-type LDB1 overexpression in flies and was unable to rescue lethality or locomotor defects in a *chi*-deficient background but instead led to even worse phenotypes. Fly experiments therefore postulate a toxic effect for the C-terminal variants, which can be explained by the dominant-negative effect observed on a cellular level. Additionally, C-terminal LID-disrupting frameshift variants resulted in increased protein expression, likely due to an impaired degradation of LDB1 due to the loss of ubiquitination site p.Lys401,^40,41^ which potentially further exacerbates the observed dominant-negative effect. Interestingly, these frameshift variants lead to a non-native extended C-terminal tail, and also resulted in the formation of small nuclear aggregates. For similar C-terminal frameshifting variants, a novel mutational mechanisms involving phase separation has recently been proposed.^42^ As nuclear punctate localization in our study differs from previous findings of nucleolar mislocalization, mutational mechanisms may be slightly different.

For two of three N-terminal missense variants located in the dimerization domain [p.(Arg121Trp) and p.(Arg193Trp)], we observed impaired homodimerization of LDB1, likely leading to a loss-of-function effect. LDB1 homodimerization is essential for gene activation by enabling long range DNA looping. For example, LDB1 activates β-globin transcription in erythroid cells by bridging the distant locus control region (LCR) enhancer and β-globin promotor. The LDB1-DD alone fused with LMO2 can fully rescue transcription in LBD1 depleted murine erythroleukemia cells.^41^ Other studies have also shown that the LDB1/chi-DD is needed for LDB1 functions, including axon guidance, motoneuronal differentiation and hippocampal development.^4,43^ In addition, the variant p.(Arg193Trp) also decreases protein expression of LDB1, possibly due to reduced protein stability as predicted by structural modeling, showcasing a second possible disease mechanism for this variant. In *Drosophila*, overexpression of LDB1 with any of these variants did not reduce viability or affect sleep patterns and could not rescue lethality or geotaxis phenotypes in a *chi*-deficient background. Therefore, both *in vitro* and *in vivo* experiments highly suggest that p.(Arg121Trp) and p.(Arg193Trp) are loss-of-function variants. We accordingly considered our experimental results sufficient evidence to apply the PS3 criterion in the ACMG criteria for variant classification for these and the C-terminal LID-affecting variants.

By contrast we did not observe any functional effects in cellular assays for the third N-terminal missense variant p.(Arg181Gln). In *Drosophila*, overexpression and rescue with mutant LDB1 with this variant in the lethality assay also did not indicate functional impairment. Rescue in the negative Geotaxis assay yielded intermediate, not fully conclusive results, indicating a possibly mild impairment of LDB1 function due to this variant. Sleep analysis however showed activity levels closer to the control than flies with an overexpression of wild-type LDB1, which would be indicative of a loss-of-function effect. As most of the other experiments did not confirm a loss-of-function effect, it remains unclear whether this variant is pathogenic or not. This variant notably has the lowest pathogenicity scores out of all the studied variants and structural modeling suggested only minor changes to the protein structure due to this variant. Interestingly, the individual harboring this variant is one of two individuals with seizures in our cohort, and the only one with a severe neonatal onset therapy-resistant epilepsy.

N-terminal LGD variants were not included in cellular functional assays as they likely undergo NMD and are likely leading to a loss-of-function mechanism. Heterozygous loss-of-function variants in LDB1 likely result in haploinsufficiency, as the gene is predicted to be dosage sensitive. Gene knockdown in *Drosophila* is an excellent model for haploinsufficiency since it reduces gene expression by around 50%. Dosage sensitivity of *LDB1/chi* was seen *in vivo* as locomotor defects upon pan-neuronal but not glial knockdown of *chi* were observed with a stronger knockdown also resulting in more severe defects, confirming deletion intolerance of *chi/LDB1* and highlighting the importance of LDB1 in the nervous system. In line with the fact that LDB1 forms a complex with LHX3 and ISL1 that is essential for the differentiation of motoneurons in chick embryos,^44^ motoneuronal knockdown of *chi* also caused locomotor defects but to a lesser degree than pan-neuronal knockdown. Sleep architecture was altered upon pan-neuronal knockdown but could be confounded by the flies’ impaired locomotor activities. Glial knockdown, however, also consistently increased the duration of sleep bouts, indicating that regulation of sleep is indeed influenced by *chi*. Sleep disturbances are among the most common co-occurring features of NDDs, affecting up to 86% of individuals (compared to ∼20% of typically developing children^45^). These disturbances substantially reduce the quality of life for both patients and their families and are often reported as a greater burden than other physical or cognitive challenges.^45^ Given our findings that the *LDB1* orthologue *chi* plays a key role in sleep regulation, it would be valuable to investigate in more detail whether subtle sleep disturbances are present in this LDB1 patient cohort and to systematically characterize their nature and severity. Our data suggest that *LDB1/chi* is important both in neurons and glia cells, but plays a more prominent role in neurons. In summary the experiments emphasize the importance of LDB1 in the brain and confirm its dosage sensitivity, making it likely that N-terminal LGD and missense variants result in haploinsufficiency.

In conclusion, we refine the spectrum of *LDB1*-associated NDDs by identifying a novel genotype-phenotype correlation with two overlapping but distinct *LDB1*-related disorders with distinct disease mechanisms. Individuals with C-terminal LID-affecting variants present with a more severe phenotype and congenital ventriculomegaly and variants act in a dominant-negative way, whereas individuals with N-terminal missense or LGD variants seem to be more mildly affected and do not share this feature, and those variants likely lead to haploinsufficiency either through loss-of-function missense mutations or NMD.

## Data Availability

Data generated or analyzed during this study are included in the manuscript and/or the corresponding supplemental information. All variants will be submitted to ClinVar (accession number: XXX). This paper does not report original code. Generated reagents (plasmids and fly lines) are available from the corresponding author upon request with a completed material transfer agreement.

## Acknowledgements

We thank the affected individuals and their families for their participation in this study. We thank Pleuni Schreurs for her excellent experimental support. A.G. is supported by a young investigator grant from the Bern Center for Precision Medicine (BCPM). This study makes use of data generated by the DECIPHER community. A full list of centres who contributed to the generation of data is available from https://deciphergenomics.org/about/stats and via email from. DECIPHER is hosted by EMBL-EBI and funding for the DECIPHER project was provided by the Wellcome Trust [grant number WT223718/Z/21/Z]. *Drosophila* stocks obtained from the Bloomington Drosophila Stock Center (NIH P40OD018537) from the Vienna Drosophila Resource Center (VDRC, www.vdrc.at). Plasmids were obtained from the Drosophila Genomics Resource Center (NIH Grant 2P40OD010949) (pUASTattb). F.N. is member of the European Reference Network for Rare Neurological Diseases - Project ID No 739510. C.Z. is supported by a grant from the Swiss National Science Foundation (SNSF, 10001220) and is a member of the European Reference Network on Rare Congenital Malformations and Rare Intellectual Disability ERN-ITHACA, funded by the European Union (grant agreement No 101156387). KM, SD and JG are members of the German Center for Child and Adolescent Health (DZKJ; project No 01GL2402A).

## Author contributions

Conceptualization: A.G.; Data curation and investigation: R.F., M.C.-T., C.Z., H.S., A.G.; Data collection: all; Formal analysis: R.F., M.C.-T., H.S., A.G.; Supervision: A.G.; Visualization: R.F., M.C.-T., H.S., A.G.; Writing – original draft: R.F., M.C-T., A.G.; Writing – review and editing: all.

